# Estimating the daily trend in the size of the COVID-19 infected population in Wuhan

**DOI:** 10.1101/2020.02.12.20022277

**Authors:** Qiushi Lin, Taojun Hu, Xiao − Hua Zhou

## Abstract

There has been an outbreak of coronavirus disease (COVID-19) in Wuhan city, Hubei province, China since December 2019. Cases have been exported to other parts of China and more than 20 countries. We provide estimates of the daily trend in the size of the epidemic in Wuhan based on detailed information of 10,940 confirmed cases outside Hubei province.

## Background

As of February 13, 2020, the National Health Commission (NHC) of China has confirmed a total of 63,851 cases of COVID-19 in mainland China, including 10,204 severe cases, 1,380 deaths, and 6,723 recoveries. An additional total of 10,109 suspected cases were reported. Wuhan, the epicenter of the COVID-19 outbreak, has 35,991 confirmed cases. The NHC has also received 53 confirmed reports in Hong Kong Special Administrative Region, China, 10 in Macau Special Administrative Region, China, and 18 in Taiwan, China. [1] More than 500 cases have been detected outside China.

Despite the considerable medical resources and personnel that have been dispensed to combat COVID-19 in Hubei province, hospital capacity continues to be overburdened. There continues to be a shortage of hospital beds needed to accommodate the rising number of COVID-19 patients. In response to this growing crisis, Wuhan plans to transform hotels, venues, training centers and college dorms into quarantine and treatment centers for COVID-19 patients. Further, 13 mobile cabin hospitals will be built to provide over 10,000 beds. [2] Therefore, a careful and precise understanding of the potential number of cases in Wuhan is crucial for the prevention and control of the COVID-19 outbreak. Wu et al. (2020) provided an estimate of the total number of cases of COVID-19 in Wuhan, using the number of cases exported from Wuhan to cities outside mainland China. [3] However, since the number of cases exported from Wuhan to cities outside mainland China is small, their estimate of the size of the epidemic in Wuhan may not be precise and has large variability. Using the number of cases exported from Wuhan to all cities, including cities in China, outside Hubei Province, You et al. (2020) proposed a new method to estimate the total number of cases of COVID-19 in Wuhan. [4] However, their method can only give an estimate of the cumulative number of cases until a certain date.

In this article, we propose a new statistical method to estimate daily number of cases in Wuhan under a similar dynamic equation model as the one in [3]. Unlike the one in [3], our method can also handle the missing information on whether a case is exported from Wuhan.

## Results

We estimate the number of cases that should be reported in Wuhan by January 11, 2020, is 4,094 (95% confidence interval [CI]: 3,980 – 4,211) and 58,153 (95% CI: 56,532 – 59,811) by February 13, 2020. Figure 1 shows how the estimated number of cases in Wuhan increases over time, together with the 95% confidence bands. As shown in Figure 2, the reporting rate has grown rapidly from 1.41% (95% CI: 1.37% - 1.45%) on January 20, 2020, to 61.89% (95% CI: 60.17% - 63.66%) on February 13, 2020. The date of first infection is estimated as November 30, 2019.

**Figure 1.**
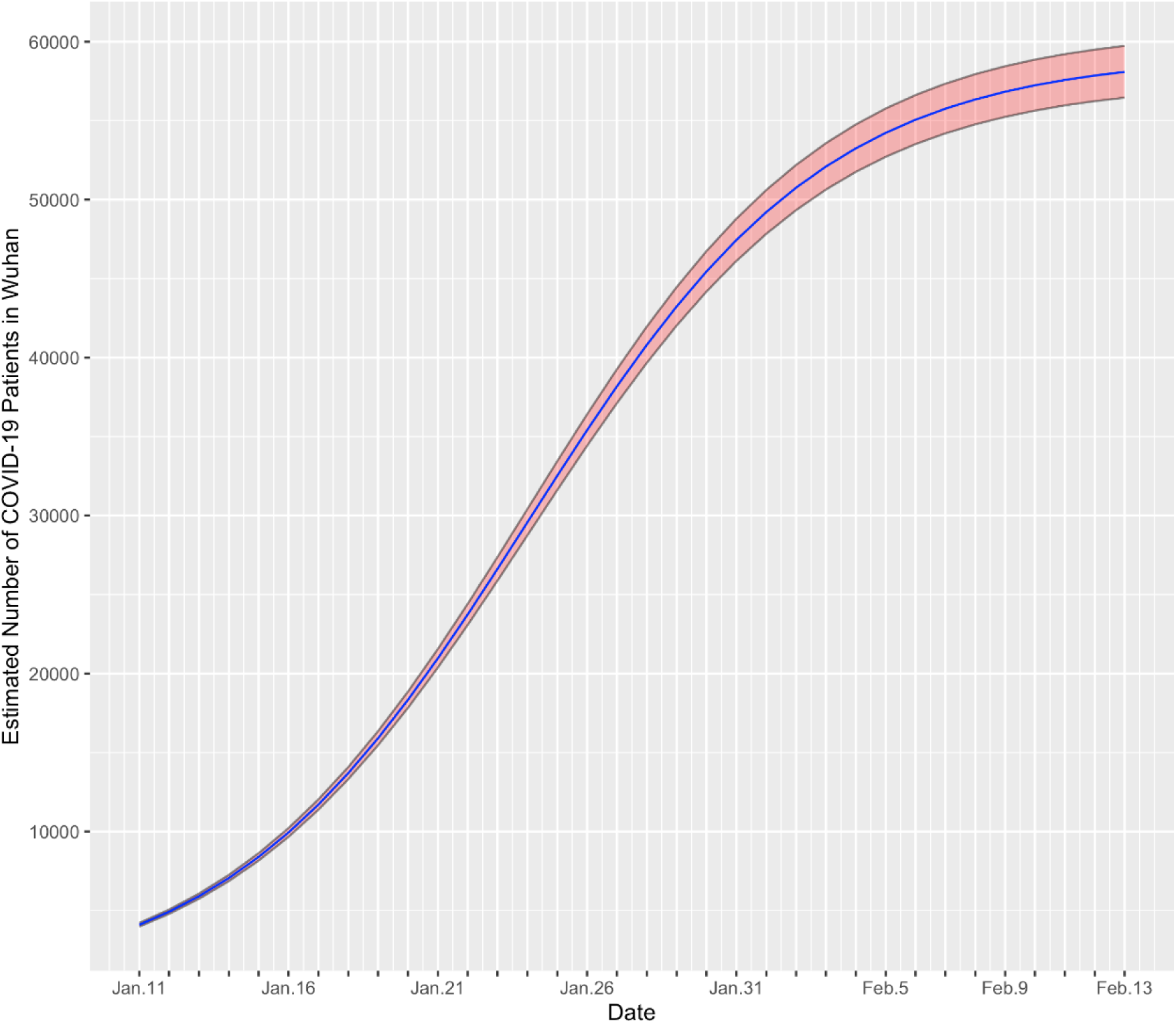
Estimated number of total cases in Wuhan.

**Figure 2.**
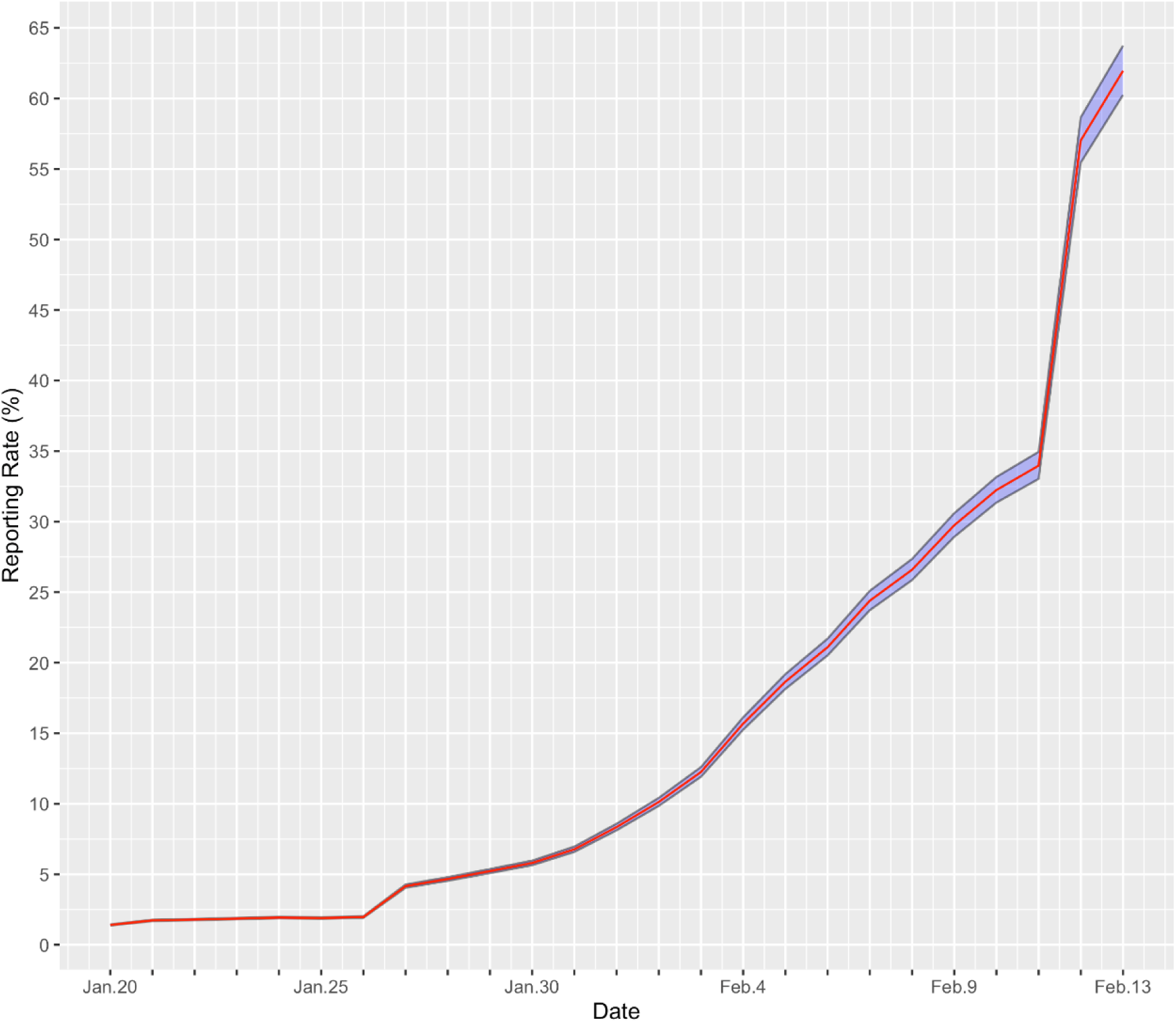
The ratio of reported number of cases to the estimated number.

## Data Description

Data retrieved from publicly available records from provincial and municipal health commissions in China and ministries of health in other countries included detailed information for 10,940 confirmed cases outside Hubei province, including region, gender, age, date of symptom onset, date of confirmation, history of travel or residency in Wuhan, and date of departure from Wuhan. Among the 7,500 patients with gender data, 3,509 (46.79%) are female. The mean age of patients is 44.48 and the median age is 44. The youngest confirmed patient outside Hubei province was only five days old while the oldest is 97 years old.

We display the epidemiological data categorized by the date of confirmation in Table 2. An imported case means a patient that had been to Wuhan and was detected outside Hubei province. A local case means a confirmed case that had not been to Wuhan. Among the total of 10,940 cases, 6,903 (63.10%) have such epidemiological information. The number of imported cases reached its peak on January 29, 2020, and the fourth column of Table 2 shows that the proportion of imported cases declines over time. This might reflect the effect of containment measures taken in Hubei province to control the COVID-19 outbreak. [5] Meanwhile, the daily counts of local cases are over 300 from February 2, 2020, to February 7, 2020, which indicate that infections among local residents should be a major concern for authorities outside Hubei province.

**Table 1.** Demographic Characteristics of Patients with COVID-19 outside Hubei Province.

| Age group<br>- years | Female<br>(N=3,509) | Male<br>(N=3,991) | No information<br>(N=3,440) |
| --- | --- | --- | --- |
| 0-20 | 97(3)* | 149(4) | 2 |
| 20-39 | 1,076(33) | 1,348(36) | 41 |
| 40-59 | 1,425(43) | 1,598(43) | 38 |
| 60-79 | 630(19) | 578(15) | 39 |
| ≥80 | 66(2) | 60(2) | 7 |
| No information | 215 | 258 | 3,313 |

\* Number (%). The percentages do not take missing data into account.

**Table 2.**
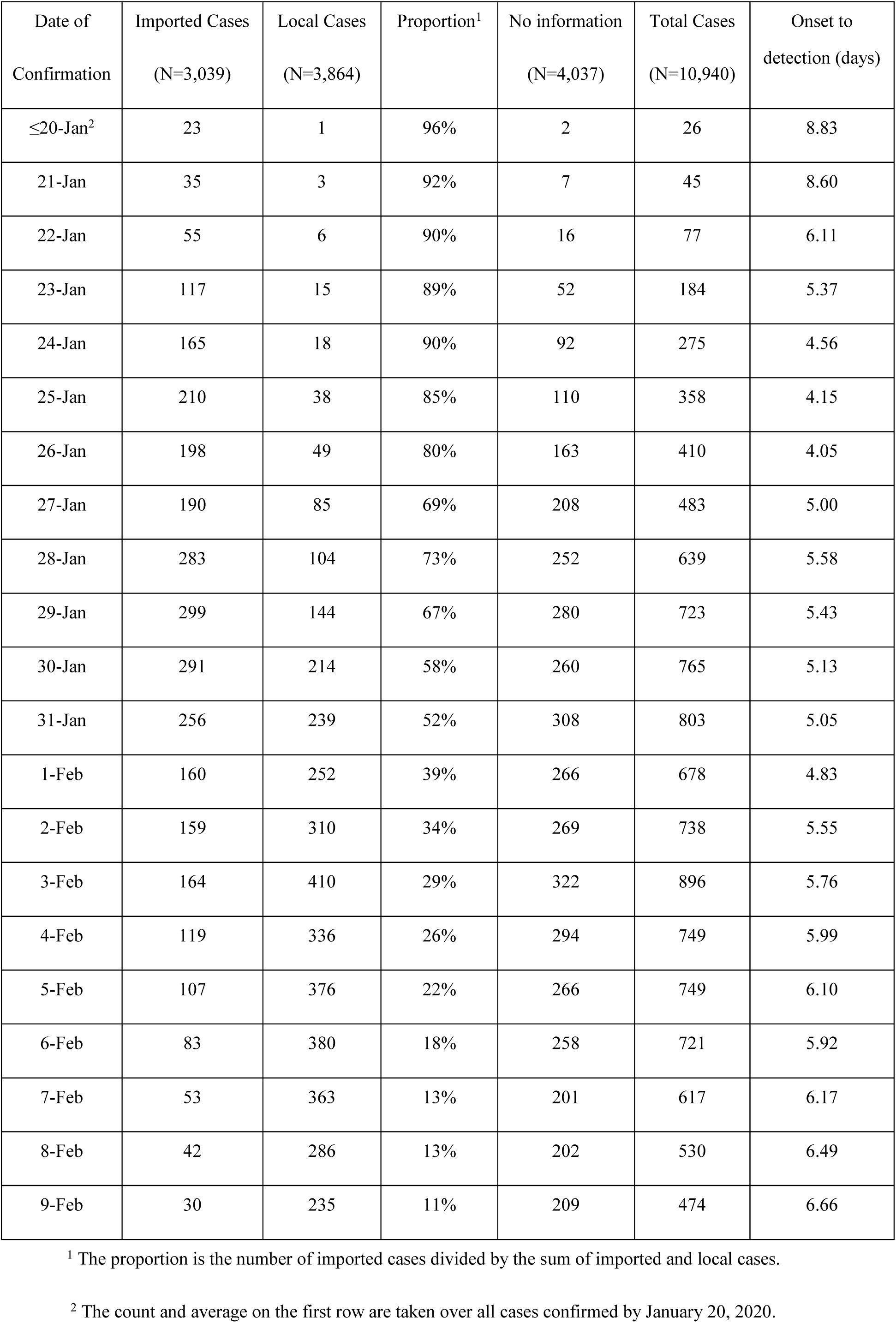
Patient data categorized by the date of confirmation.

| Date of Confirmation | Imported Cases<br>(N=3,039) | Local Cases<br>(N=3,864) | Proportion<sup>1</sup> | No information<br>(N=4,037) | Total Cases<br>(N=10,940) | Onset to detection (days) |
| --- | --- | --- | --- | --- | --- | --- |
| ≤20-Jan<sup>2</sup> | 23 | 1 | 96% | 2 | 26 | 8.83 |
| 21-Jan | 35 | 3 | 92% | 7 | 45 | 8.60 |
| 22-Jan | 55 | 6 | 90% | 16 | 77 | 6.11 |
| 23-Jan | 117 | 15 | 89% | 52 | 184 | 5.37 |
| 24-Jan | 165 | 18 | 90% | 92 | 275 | 4.56 |
| 25-Jan | 210 | 38 | 85% | 110 | 358 | 4.15 |
| 26-Jan | 198 | 49 | 80% | 163 | 410 | 4.05 |
| 27-Jan | 190 | 85 | 69% | 208 | 483 | 5.00 |
| 28-Jan | 283 | 104 | 73% | 252 | 639 | 5.58 |
| 29-Jan | 299 | 144 | 67% | 280 | 723 | 5.43 |
| 30-Jan | 291 | 214 | 58% | 260 | 765 | 5.13 |
| 31-Jan | 256 | 239 | 52% | 308 | 803 | 5.05 |
| 1-Feb | 160 | 252 | 39% | 266 | 678 | 4.83 |
| 2-Feb | 159 | 310 | 34% | 269 | 738 | 5.55 |
| 3-Feb | 164 | 410 | 29% | 322 | 896 | 5.76 |
| 4-Feb | 119 | 336 | 26% | 294 | 749 | 5.99 |
| 5-Feb | 107 | 376 | 22% | 266 | 749 | 6.10 |
| 6-Feb | 83 | 380 | 18% | 258 | 721 | 5.92 |
| 7-Feb | 53 | 363 | 13% | 201 | 617 | 6.17 |
| 8-Feb | 42 | 286 | 13% | 202 | 530 | 6.49 |
| 9-Feb | 30 | 235 | 11% | 209 | 474 | 6.66 |

<sup>1</sup> The proportion is the number of imported cases divided by the sum of imported and local cases.
<sup>2</sup> The count and average on the first row are taken over all cases confirmed by January 20, 2020.

The last column of Table 2 lists the mean time from symptom onset to confirmation for patients confirmed on each day. The median duration of all cases is 5 days, and the mean is 5.54 days. In general, the detection period decreased in the first week after January 20, 2020, but increased since then. The improvements in detection speed and capacity might cause the initial decline, and the rise may be due to more thorough screening, leading to the detection of patients with mild symptoms who would otherwise not go to the hospitals. [6]

### Assumptions

The proposed method relies on the following assumptions:

1. Between January 10, 2020, and January 23, 2020, the average daily proportion of departing from Wuhan is *p*.
2. There is a *d* = *d*_1_ + *d*_2_-day window between infection to detection, including a *d*_1_-day incubation period and a *d*_2_-day delay from symptom onset to detection.
3. Trip durations are long enough that a traveling patient infected in Wuhan will develop symptoms and be detected in other places rather than after returning to Wuhan.
4. All travelers leaving Wuhan, including transfer passengers, have the same risk of infection as local residents.
5. Traveling is independent of the exposure risk to COVID-19 or of infection status.
6. Patients are not able to travel *d* days after infection.
7. Recoveries are not considered in this method.
8. The proportion of imported cases in the patients with no information is the same as the observed proportion on each day.

We next make some remarks about our assumptions.

a. January 10, 2020, is the start of Chinese New Year travel rush, and January 23, 2020, is the date of Wuhan lockdown. [5] In the total of 10,940 cases, only 131 (1.2%) cases’ date of departure from Wuhan are not in this period. They are excluded from our analysis.
b. If the true average daily proportion of leaving Wuhan is larger the assumed *p*, this violation of Assumption 1 could lead to overestimation of the number of cases in Wuhan,
c. If the average time from infection to detection is longer than the assumed *d* days, this violation of Assumption 2 would lead to an overestimation.
d. If travelers have a lower risk of infection than residents in Wuhan, this violation of Assumption 4 would cause an underestimation.
e. If infected individuals are less likely to travel due to the health conditions, this violation of Assumption 5 would cause an underestimation.
f. Given that the number of recoveries in early days of outbreak is relatively small compared to the number of COVID-19 patients, Assumption 8 should not significantly influence the result.

We perform Sensitivity Analysis on the effect of some of the violations on our results.

## Methods

The spread of COVID-19 outside Hubei province is relatively controlled given the adequate medical resources. We use the reported number outside Hubei as it is a fairly accurate representation of the actual epidemic situation. In this modelling study, we first estimate the epidemic size in Wuhan from January 11, 2020, to February 13, 2020, based on the confirmed cases outside Hubei province that left Wuhan by January 23, 2020. Since some confirmed cases have no information on whether they visited Wuhan before, we adjust the number of imported cases after taking these missing values into account. We then calculate the reporting rate in Wuhan from January 20, 2020, to February 13, 2020. Finally, we estimate the date when the first patient was infected.

### Notations

Let Day *t*_0_ denote the date of infection for the very first case. Let *N*_*t*_ be the cumulative number of cases that should be confirmed in Wuhan by Day *t*. Other notations of our model are defined in Table 3.

**Table 3.** Notations for our model.

|  |  |
| --- | --- |
| $T_t$ | The reported number of confirmed cases outside Hubei province on Day $t$ |
| $I_t$ | The observed number of imported cases on Day $t$ |
| $L_t$ | The observed number of local cases on Day $t$ |
| $U_t$ | The number of cases with no information on Day $t$, $U_t = T_t - I_t - L_t$ |
| $x_t$ | The number of imported cases on Day $t$, $x_t$ is unknown. |
| $X_t$ | The cumulative number of imported cases by Day $t$, $X_t = \sum_{k=1}^t x_k$ |
| $N_t$ | The cumulative number of cases that should be confirmed in Wuhan by Day $t$ |
| $t_0$ | The date of infection for the very first case |
| $p$ | The daily probability of departing from Wuhan |
| $d$ | The time from infection to detection. |
| $t_c$ | The time when $N_t$ increases at the fastest rate. |
| $r$ | A parameter that determines the growth rate of $N_t$ |
| $K$ | The size of the population that are susceptible to COVID-19 in Wuhan. |

The numbers $T_t$ , $I_t$ , and $L_t$ are the observed data used in our model, $t_c$ , $r$ , and $K$ are the parameters that determine how $N_t$ changes over time.

### Model

The growth trend of the size *N*_*t*_ of infected population is determined by the following ordinary differential equation:

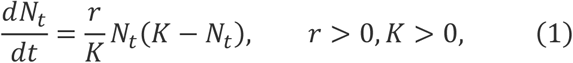

where *K* is the size of the population that are susceptible to COVID-19 in Wuhan, and r is a constant that controls the growth rate of *N*_*t*_. This is the simplified version of the famous SIR model [3, 7] in epidemiology. It is a good model at early stage of the epidemic when the number of recoveries is still relatively small compared to infected cases. The growth rate of *N*_*t*_ is proportional to the product of *N*_*t*_ and the number *K* − *N*_*t*_ of people that are susceptible but not infected yet. The equation (1) has an analytical solution

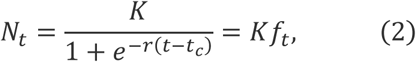

Where 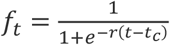, and the derivative 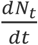 is maximized at 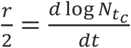 is the growth rate of log *N*_*t*_ at time *t*_*c*_, *K* is a parameter to be estimated.

### Estimation

We use data on the confirmed cases who left Wuhan between January 10, 2020 and January 23, 2020, to estimate *K*. Under Assumption 2, cases infected on Day *t* will be detected on Day *t* + d, so the number of infected cases in Wuhan is *N*_*t+d*_ on Day *t*. If *t*_0_ ≤ *t* ≤ *t*_0_ + d, there should be no confirmed cases. If *t*_0_ + *d* < *t* ≤ *t*_0_ + 2d, imported cases on Day *t* are infected in Wuhan on Day *t* − d. If *t* > *t*_0_ + 2d, under Assumption 6, N_*t*−*d*_ patients are not able to travel. There are *N*_*t*_ infected cases in Wuhan on Day *t* − *d*, the number of imported cases *x*_*t*_ on Day *t* follows a binomial(*N*_*t*_, *p*) distribution, where p is the assumed average daily proportion of leaving Wuhan between January 10, 2020, and January 23, 2020. Let *X*_*t*_ be the cumulative number of imported cases by Day *t*, then

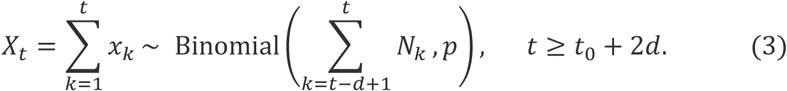

From equations (2) and (3), 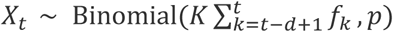. The parameter estimate 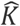 is derived by maximizing the likelihood function

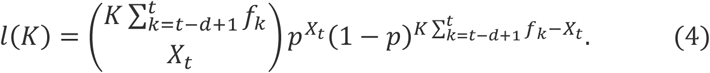

The lower and upper bound of the 95% confidence interval 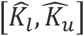 are values such that the cumulative distribution function 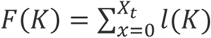 equals 0.975 and 0.025, respectively. The reporting rate is the reported cumulative number of cases in Wuhan on Day t divided by our estimated number 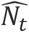. The estimate of the date *t*_0_ of first infection is obtained by solving the equation 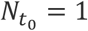.

Determining the number of imported cases *x*_*t*_ plays a crucial role in the modeling procedure. Note that not all cases have clear records on the history of travel or residency in Wuhan, we need to impute the missing values. Under Assumption 8, the proportion of imported cases in the *U*_*t*_ patients with no information is the same as the observed proportion 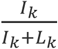. Therefore,

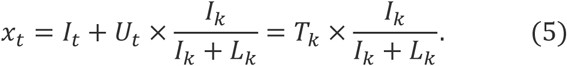

The average daily proportion of leaving Wuhan between January 10, 2020 and January 23, 2020 is estimated to be the ratio of daily volume of travelers to the population of Wuhan (14 million). More than 5 million people have left Wuhan due to the Spring Festival and epidemic. [8]. The Chinese New Year travel rush started at January 10, 2020, and the lockdown of Wuhan city happened on January 23, 2020. During the travel rush, 34% of the passengers traveled across 300 km. [9] Major cities outside Hubei province are generally over 300 km from Wuhan. This would imply, on average, the daily probability p of traveling from Wuhan to places outside Hubei province would be 5*0.34/14/14=0.009. Li et al. estimated that the mean incubation period of 425 patients with COVID-19 was 5.2 days (95% CI, 4.1 - 7.0). [10] The mean time from symptom onset to detection calculated from our data is 5.54 days, so we choose *d*_1_ = *d*_2_ = 5 days. January 29, 2020, has the maximum count of imported cases. Since *x*_*t*_ has a binomial(*N*_*t*_ − *N*_*t−d*_, *p*) distribution with constant *p, N*_*t*_ − *N*_*t−d*_ also reaches its maximum at *t* = January 29, 2020. From the logistic function (2), *t*_*c*_ is the midpoint of *t* and *t* − *d*, that is 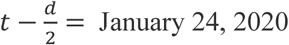, which is shortly after the lockdown of Wuhan city. [5] Wu et al. estimated the epidemic doubling time as 6.4 days (95% CI: 5.8 – 7.1) as of January 25, 2020. [3] From this result, we estimate that 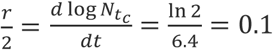. Using these values for parameters *p, d, t*, and *r*, we can derive the maximum likelihood estimate 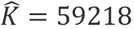, with 95% confidence interval [57567, 60906].

### Sensitivity Analysis

We explore the sensitivity of the estimate of total cases in Wuhan to our assumptions and choices of parameters *p, d*, and *r*. Note that 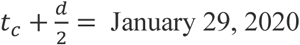.

Compared to the baseline, the parameters are expanded or shrunk by about 30% to reflect the possible uncertainty. Table 4 summaries the estimate the number of cases should be reported on January 11, 2020, and February 13, 2020, under baseline assumptions and alternative scenarios. Confidence intervals are omitted. The currently reported number 35,991 on February 13, 2020, is substantially smaller than the estimate of our most conservative scenario.

**Table 4.** Estimated case numbers on January 11 and February 13, 2020, based on different choices of parameters.

|  | Baseline | Scenario 1 | Scenario 2 | Scenario 3 | Scenario 4 | Scenario 5 | Scenario 6 |
| --- | --- | --- | --- | --- | --- | --- | --- |
| $p$ | 0.009 | 0.012 | 0.006 | 0.009 | 0.009 | 0.009 | 0.009 |
| $d = d_1 + d_2$ | 10 | 10 | 10 | 13 | 7 | 10 | 10 |
| $r$ | 0.2 | 0.2 | 0.2 | 0.2 | 0.2 | 0.25 | 0.15 |
| 11-Jan | 4,094 | 3,071 | 6,141 | 3,985 | 4,724 | 2,123 | 7,877 |
| 13-Feb | 58,153 | 43,614 | 87,228 | 43,409 | 86,675 | 56,497 | 60,243 |

## Conclusions

The estimated reporting case rate has increased rapidly, reaching over 30% by February 11, 2020. It is almost doubled in the following two days, mainly due to the inclusion of 14,031 clinically diagnosed cases in the case reports of Wuhan. This might indicate that the testing capacity of Wuhan is insufficient. Clinical diagnosis could be a good complement to the current method of confirmation. The currently reported number of 35,991 cases as of February 13, 2020, is still far below our estimate of 58,153. There may still be a lot of unreported cases. More thorough screening of all patients with a mild or moderate symptoms of respiratory diseases should be conducted to better control the spread of COVID-19.

## Data Availability

Data retrieved from publicly available records from provincial and municipal health commissions in China and ministries of health in other countries included detailed information for 10,940 confirmed cases outside Hubei province, including region, gender, age, date of symptom onset, date of confirmation, history of travel or residency in Wuhan, and date of departure from Wuhan.

## References

[1] National Health Commission Update on February 10, 2020. National Health Commission of the People’s Republic of China. http://weekly.chinacdc.cn/news/TrackingtheEpidemic.htm#NHCFeb14 [2020-2-14]

[2] Hubei ordered to admit all patients in hospitals. China Daily. https://www.chinadaily.com.cn/a/202002/09/WS5e3fba1ca3101282172760aa.html [2020-2-9]

[3] Wu, Jianhong & Leung, Kathy & Leung, Gabriel. (2020). Nowcasting and forecasting the potential domestic and international spread of the 2019-nCoV outbreak originating in Wuhan, China: a modelling study. The Lancet. 10.1016/S0140-6736(20)30260-9.

[4] Chong You, Qiushi Lin, Xiao-hua Zhou. An Estimation of the Total Number of Cases of NCIP (2019-nCoV) — Wuhan, Hubei Province, 2019–2020[J]. China CDC Weekly, 2020, 2(6): 87–91.

[5] China declares lockdown in Wuhan on Thursday due to coronavirus outbreak. Tass. https://tass.com/world/1111981 [2020-1-23]

[6] Beijing to set up checkpoints in all residential communities. China Daily. https://www.chinadaily.com.cn/a/202002/10/WS5e415cb1a3101282172766c4.html [2020-2-1]

[7] Kermack WO, McKendrick AG. A Contribution to the Mathematical Theory of Epidemics”. Proceedings of the Royal Society A, 1927, 115 (772): 700–721.

[8] million-plus leave Wuhan: Mayor. China Daily. https://www.chinadaily.com.cn/a/202001/27/WS5e2dcd01a310128217273551.html [2020-1-27]

[9] Big data perspective: Wuhan in the Chinese New Year travel rush. Daily Economic News. https://m.nbd.com.cn/articles/2020-01-22/1402239.html [2020-1-22] (In Chinese).

[10] Qun Li, et al. Early Transmission Dynamics in Wuhan, China, of Novel Coronavirus– Infected Pneumonia. N Engl J Med. 2020 January 29.

